# High-sensitivity cardiac troponin I and risk of dementia: 25-year longitudinal study in the Whitehall II cohort

**DOI:** 10.1101/2024.11.19.24317589

**Authors:** Yuntao Chen, Martin Shipley, Atul Anand, Dorien M Kimenai, Klaus P Ebmeier, Severine Sabia, Archana Singh-Manoux, John Deanfield, Mika Kivimaki, Gill Livingston, Nicholas L Mills, Eric J Brunner

**Affiliations:** Division of Psychiatry, University College London, London, UK; Department of Epidemiology and Public Health, University College London, London, UK; British Heart Foundation Centre for Cardiovascular Science, University of Edinburgh, Edinburgh, UK; Department of Psychiatry and Wellcome Centre for integrative Neuroimaging, University of Oxford, Oxford, UK; Inserm U1153, Epidemiology of Ageing and Neurodegenerative diseases, Universite de Paris, Paris, France; Institute of Cardiovascular Science, University College London, London, UK; Usher Institute, University of Edinburgh, Edinburgh, UK

## Abstract

**Objective:** We hypothesise that subclinical myocardial injury during midlife, indexed by increases in cardiac troponin I, is associated with accelerated cognitive decline, smaller structural brain volume, and higher risk of dementia.

**Design:** Longitudinal cohort study

**Setting:** Civil service departments in London (Whitehall II study)

**Participants:** 5985 participants aged 45-69 had cardiac troponin I measured by high-sensitivity assay at baseline (1997-99) for prospective cohort analyses. A nested case-control sample of 3475 participants (695 dementia cases and 2780 matched controls) was used for backward cardiac troponin I trajectory analysis. 641 participants provided magnetic resonance imaging (MRI) scans for brain volume analysis.

**Main outcome measures:** Incident dementia cases were ascertained from national hospital episode statistics, mental health and mortality registers until 2023. Cognitive testing was performed at six waves over 25 years (1997-99, 2002-04, 2007-09, 2012-13, 2015-16, 2019-22). Brain volume metrics were derived from structural MRI scans (2012-16).

**Results:** For prospective cohort analyses, 606 (10.1%) incident cases of dementia were recorded over a median follow-up of 24.8 years. Doubling of cardiac troponin was associated with 11% higher risk of dementia (HR=1.11, 95% CI: 1.04 to 1.19). Participants with increased cardiac troponin at baseline had a faster decline of cognitive function with age. Compared to participants with concentrations below the limit of quantitation (<2.5 ng/L), those in the upper third (>5.2 ng/L) had similar global cognitive z score at age 60, but had 0.10 (95% CI: 0.02 to 0.18) standard deviations lower score at age 80, and 0.19 (0.03 to 0.35) standard deviations lower score at age 90. Participants with dementia had increased cardiac troponin concentrations compared with those without dementia between 7 and 25 years before diagnosis. Compared to those with low cardiac troponin level (<2.5 ng/L at baseline) those with concentrations >5.2 ng/L had lower grey matter volume and higher hippocampal atrophy 15 years later, equivalent to ageing effects of 2.7 and 3 years, respectively.

**Conclusions:** Subclinical myocardial injury at midlife was associated with higher dementia risk in later life.

**What is already known on this topic:** A systematic review of observational studies suggests higher cardiac troponin concentrations are associated with poorer cognitive function and increased dementia risk. Formal meta-analysis was not performed due to the small number of available studies.

Existing studies assessed cardiac troponin once and had relatively short follow-up time. Evidence is lacking on the time course of cardiac troponin level before diagnosis in dementia cases compared with controls.

**What this study adds:** People with increased cardiac troponin I concentrations in mid-life had faster cognitive decline and were more likely to develop dementia over 25 years of follow-up. Backward trajectory analysis using three measurements using a high-sensitivity assay showed that people with dementia had higher cardiac troponin levels as early as 25 years before dementia diagnosis compared with those without dementia. People in the magnetic resonance imaging subcohort with higher cardiac troponin I concentrations at baseline had lower grey matter volume and hippocampal atrophy 15 years later.

Subclinical myocardial injury in midlife, by indicating long-term risk of dementia, is unlikely to be due to preclinical changes before dementia onset and may lie on the causal pathway to dementia.

## Introduction

Emerging evidence suggests an intertwined relation between the health of heart and brain.^1–3^ The 2024 Lancet Commission on dementia suggests 17% of dementia cases can be prevented or delayed by modifying cardiovascular risk factors including high cholesterol, physical inactivity, diabetes, hypertension, obesity, smoking and excessive alcohol consumption.^4^ People with poor cardiovascular health, even in the absence of clinical coronary heart disease, have increased risk of dementia.^5,6^

Cardiac troponin is a commonly-used cardiac biomarker and its utility has been extended with introduction of high-sensitivity assays.^7–9^ Modest elevation in cardiac troponin in apparently healthy populations is linked with increased risk of dementia, although existing studies have only assessed cardiac troponin once and had relatively short follow-ups.^10–12^ Consequently, it is unknown how long before dementia diagnosis cardiac troponin levels become increased among cases relative to controls. Further, few studies have prospectively examined whether cardiac troponin in midlife is a marker of potential preclinical changes in dementia-related measures such as cognition and brain volume. Consistent results from these different lines of research would strengthen the evidence on the role of subclinical myocardial injury in the aetiology of dementia.

This study uses longitudinal data from the Whitehall II cohort study spanning 25 years and three measurements of cardiac troponin I based on a high-sensitivity assay. We examined 1) the long-term association of cardiac troponin I at study baseline with cognitive trajectory and incident dementia in prospective analyses; 2) how long before diagnosis of dementia the cardiac troponin I level has been elevated by modelling the backward trajectory of cardiac troponin I; and 3) the association of cardiac troponin I with brain volume measures assessed 15 years later in a substudy of structural magnetic resonance imaging.

## Methods

### Study population

The Whitehall II study is an ongoing longitudinal study of 10 308 people recruited from the British Civil Service in 1985-88. Participants had clinical examinations of behavioural and biomedical factors in 8 waves (1985-88, 1991-94, 1997-99, 2002-04, 2007-09, 2012-13, 2015-16, 2019-22). Written informed consent from participants was renewed at each contact. The baseline for this study is 1997-99 when cardiac troponin was first measured. We included 6 waves of clinical examination until 2022. Disease and death data up to March 2023 were obtained through linkage to electronic health records of the UK National Health Service.

## Measures

### High-sensitivity cardiac troponin I

Cardiac troponin I was measured using stored blood samples at three waves (1997-99, 2007-09, 2012-13).^9,13^ Blood samples for each wave were handled according to a standardised protocol. Fasting venous blood samples were collected, centrifuged, and serum was stored in aliquots at -80 °C until batch analysis was performed. Cardiac troponin I concentrations were measured using the Siemens Atellica Immunoassay High Sensitivity Troponin I assay (Siemens Healthineers, Erlangen, Germany). This assay has a limit of blank of 0.5 ng/L, limit of detection of 1.6 ng/L, and a limit of quantitation of 2.5 ng/L. In the analyses, we assigned 0.5 ng/L to those with cardiac troponin concentration below the limit of blank. The sex-specific 99^th^ centile upper reference limits in the general population used for clinical referral are 34 ng/L and 53 ng/L in females and males, respectively.

### Cognitive function

The cognitive test battery was introduced to the Whitehall II clinic in 1997-99 and repeated at all subsequent clinical assessments. We included all six waves of cognition data between 1997 (aged 45-69 years) and 2022 (aged 68-92 years). The tests have good test-retest reliability (range 0.6-0.9), assessed in 556 participants and retested within three months in 1997-99. Three cognitive domains (memory, executive function, and fluency) were assessed. Memory was assessed using a 20-word free recall test. Executive function was assessed with the Alice Heim 4-I test. Fluency was assessed using measures of phonemic and semantic fluency. Participant were asked to recall in writing as many words beginning with “s” (phonemic fluency) and as many animal names (semantic fluency) as they could. One minute was allowed for each test.

Based on three cognitive domains, we created a global cognitive score incorporating all tests described above by firstly using the distribution of the first wave of cognitive data (1997-99) to standardise the raw scores for each domain to z scores. We summed these z scores and restandardised them to yield the global score, an approach that minimises measurement error inherent individual tests.^14^

### Dementia

We used three registers (the national hospital episode statistics (HES) database, the Mental Health Service Data Set, and the mortality register) to ascertain dementia using ICD-10 codes F00-F03, F05.1, G30, and G31. Record linkage was available up to 1 March 2023. Date of dementia was set as the earliest record of a diagnostic code for dementia in any of the three ascertainment databases. As our outcome was incident dementia, participants with dementia at baseline were excluded.

### Brain volume

771 participants, randomly selected from Whitehall II phase 11 (2012-13), received multimodal brain magnetic resonance imaging (MRI) scans at the Oxford Centre for Functional MRI of the Brain, as part of the Whitehall II imaging substudy.^15^ Due to a scanner upgrade, two MRI scanners were used: a 3T Siemens Magnetom Verio scanner (Erlangen, Germany) with a 32-channel head coil (April 2012 to December 2014) and a 3T Siemens Prisma Scanner (Erlangen, Germany) with a 64-channel head-neck coil in the same centre (July 2015 to December 2016). The scan parameters were identical or closely matched between scanners. Analyses were adjusted for scanner type. Brain tissues were segmented using an automated segmentation tool, which extracts measures of total grey matter, white matter, and cerebrospinal fluid.^15^ Intracranial volume was calculated as the sum of total grey matter, white matter, and cerebrospinal fluid. All volumes were normalised by converting them to percentages of the intracranial volume. Burden of white matter hyperintensities was estimated using the Fazekas visual rating scale.^16^ A score (range 0-3) was assigned for deep white matter and periventricular areas based on the size, location, and confluence of lesions. We used the sum of the two scores to estimate the total burden of white matter intensities. Hippocampal atrophy was estimated using the Scheltens score assessed by three clinicians independently.^17^

### Clinical characteristics

We included sociodemographic, health behaviours, and health status at baseline (1997-99) as covariates: age, sex, education (less than secondary school, secondary school, university), occupational position based on British Civil Service employment grade (administrative grades, professional or executive grades, clerical or support grades), birth cohort defined based on sociohistorical events^18^ (Depression-era cohort born 1930-38, World War 2 cohort born 1939-45, Post-war cohort born 1946-52); smoking status (never, former, current), alcohol consumption (non-drinker, moderate consumption: 1-14 units/week in women or 1-21 units/week in men, heavy consumption: >14 units/week in women or >21 units/week in men), physical activity (hours of moderate or vigorous exercise/week, categorised as low <1 hour, moderate 1-2.4 hour, high ≥2.5 hour); body mass index (BMI, kg/m^2^), systolic blood pressure (SBP, mmHg), diastolic blood pressure (DBP, mmHg), fast glucose (mmol/l), total cholesterol (mmol/L), diabetes (doctor-diagnosed diabetes or use of diabetes medication). Cardiovascular disease was identified via linkage to national hospital episode statistics (HES) data, recording any diagnosis of coronary heart disease (ICD-9 codes 410-414 and 429; ICD-10 codes I20-I25) or stroke (ICD-10 codes I60-I64 and G45).

### Statistical analysis

#### Association between cardiac troponin I at baseline and incidence of dementia

We included all participants free of dementia or cardiovascular disease, who had data on cardiac troponin I at baseline (1997-99). These participants were followed-up to date of record of dementia, death, or 1 March 2023, whichever came first. Cox proportional hazard models were used to examine the association between cardiac troponin I and dementia incidence. We modelled cardiac troponin I both continuously and categorically. For the continuous analysis, we used log2 transformation as the cardiac troponin levels were right-skewed. For the categorical analysis, we used the following categories: <2.5 ng/L (below the limit of quantitation threshold, reference group), 2.5-3.4 ng/L, 3.5-5.2 ng/L, >5.2 ng/L. The latter three groups were cut based on tertiles of troponin measurements above the limit of quantitation threshold. We also modelled cardiac troponin I using natural cubic spline function to visualise possible non-linear association with dementia incidence. In the above analyses, we used age as time scale and adjusted for sex, ethnicity, education level, occupational position, alcohol consumption, smoking status, physical activity, BMI, SBP, DBP, glucose, and total cholesterol at baseline.

#### Association between cardiac troponin I at baseline and cognitive trajectory

We included all participants that were free of dementia and cardiovascular disease at baseline, had data on cardiac troponin I at baseline, and had at least one measurement of cognitive function among 6 repeated measurements between 1997 and 2022. Mixed models with age as the time scale were used to examine whether cardiac troponin I at baseline was associated with cognitive trajectories. The model included sex, ethnicity, education level, occupational position, birth cohort, cardiac troponin I (log_2_ transformed), age, age^2^, and interactions of all the covariates with age and age^2^. A likelihood ratio test was conducted for the interaction of cardiac troponin I with age and age^2^ to examine whether the cognitive trajectory differed in different cardiac troponin I levels at baseline. We replicated the analysis using categorised cardiac troponin I in 4 groups as above to relax the linear assumption and to visualise and present the results. We plotted the cognitive trajectory by different baseline cardiac troponin level, and statistically compared cognitive scores for different troponin I groups at age 60, 70, 80, and 90. We repeated the analysis by further accounting for alcohol consumption, smoking status, physical activity, BMI, SBP, DBP, glucose, and total cholesterol.

#### Backward trajectory of cardiac troponin I before dementia

We included all participants who were free of dementia and cardiovascular disease at baseline and had at least one cardiac troponin measurement before end of follow-up. We used a nested case-control design to compare the trajectory of cardiac troponin I in those who developed dementia and those did not. Each dementia case was matched to 4 controls at the diagnostic date. We used random sampling with replacement (i.e., same control is possible to be matched with different cases). The eligibility of the control sample is: 1) free of dementia at the diagnostic date; 3) of similar sex and age (±3 years); 4) of same education level. In the sensitivity analysis, we further matched with same cardiovascular disease to see whether the effect was attenuated.

We used retrospective time since the matching date as the time scale. Time 0 was date of dementia diagnosis for each quintet (1 case and 4 matched controls). A latent process mixed model^19^ was used to model the trajectory of cardiac troponin I (log2 transformed). The model included time, time^2^, index age (age at time 0), sex, education level, case indicator (coded as 1 for cases and 0 for controls), and interactions of all the covariates (index age, sex, education level, and case indicator) with time and time^2^. Wald test was used for the interaction of case indicator with time and time^2^ to examine whether the trajectory of cardiac troponin I differed between cases and controls. We also used the Wald tests to compare the differences of predicted mean cardiac troponin I levels between cases and controls at different time points before dementia diagnosis.

#### Association between cardiac troponin I at baseline and brain volume

We included participants that were free of dementia and cardiovascular disease, did not have missing data for cardiac troponin I at phase 5 (1997-99), and had MRI scans between 2012-16. Linear regression models were used to examine the association between cardiac troponin I at baseline and whole brain volume (sum of grey and white matter), grey matter volume, and white matter volume. Poisson regression models were used to examine the association of baseline cardiac troponin I with white matter hyperintensity burden and hippocampal atrophy. We modelled cardiac troponin I both continuously and categorically aforementioned. The models were all adjusted for age at MRI assessment, sex, education, occupational position, smoking status, and alcohol consumption.

We used simple imputation for the covariates with missing values. All analyses were performed using R statistical software, version 4.4.1.

### Patient and public involvement

Participants of the Whitehall II study were not involved in setting the research question or the outcome measures, nor in developing plans for recruitment, design, or implementation of the study. No participants were asked for advice on interpretation or writing up of results. However, all results are disseminated to study participants via newsletters and our website, which has a participant portal (https://www.ucl.ac.uk/psychiatry/research/mental-health-older-people/whitehall-ii/participants-area).

## Results

7299 participants had at least one measure of cardiac troponin I. They did not have dementia at baseline. After excluding participants with cardiovascular disease (n=62) at baseline, 7237 participants were included. Among these participants, 5985 had cardiac troponin at baseline. 606 (10.1%) cases of dementia were recorded over a median follow-up time of 24.8 (interquartile range 23.3-25.1) years. Baseline characteristics, as well as by cardiac troponin I levels and dementia status at follow-up, are presented in Table 1. Participants with higher cardiac troponin I were older, more likely to be men and from higher educational and occupational group. They had higher BMI, SBP, DBP, and glucose level. Compared with those who were dementia free at the end of follow-up, people with incident dementia were older, more likely to be women, and from lower education and occupational group. They had higher BMI, glucose, SBP and total cholesterol level.

**Table 1.** Characteristics of the study sample.

|  | High-sensitivity cardiac troponin I at baseline |  | Dementia status at the end of follow-up |  |  |
| --- | --- | --- | --- | --- | --- |
|  | ≤3.4 ng/L (n=3417) | >3.4 ng/L (n=2568) | No dementia (n=5379) | Dementia (n=606) | Total population (n=5985) |
| **Age, years** | 55.3 (5.8) | 57.5 (6.1) | 55.7 (5.9) | 61.1 (5.1) | 56.2 (6.0) |
| **Women** | 1351 (39.5) | 398 (15.5) | 1539 (28.6) | 210 (34.7) | 1749 (29.2) |
| **Ethnicity, White** | 3142 (92.0) | 2350 (91.5) | 4960 (92.2) | 532 (87.8) | 5492 (91.8) |
| **Education** |  |  |  |  |  |
| Less than secondary school | 1277 (37.4) | 852 (33.2) | 1847 (34.3) | 282 (46.5) | 2129 (35.6) |
| Secondary school | 890 (26.0) | 701 (27.3) | 1455 (27.0) | 136 (22.4) | 1591 (26.6) |
| University | 1189 (34.8) | 958 (37.3) | 1971 (36.6) | 176 (29.0) | 2147 (35.9) |
| **Occupation position** |  |  |  |  |  |
| Low | 520 (15.2) | 260 (10.1) | 646 (12.0) | 134 (22.1) | 780 (13.0) |
| Intermediate | 1506 (44.1) | 1036 (40.3) | 2308 (42.9) | 234 (38.6) | 2542 (42.5) |
| High | 1347 (39.4) | 1245 (48.5) | 2361 (43.9) | 231 (38.1) | 2592 (43.3) |
| **Alcohol consumption** |  |  |  |  |  |
| Non-drinkers | 556 (16.3) | 331 (12.9) | 757 (14.1) | 130 (21.5) | 887 (14.8) |
| Moderate | 1960 (57.4) | 1539 (59.9) | 3175 (59.0) | 324 (53.5) | 3499 (58.5) |
| heavy | 788 (23.1) | 617 (24.0) | 1271 (23.6) | 134 (22.1) | 1405 (23.5) |
| **Smoking status** |  |  |  |  |  |
| Never | 1679 (49.1) | 1264 (49.2) | 2644 (49.2) | 299 (49.3) | 2943 (49.2) |
| Former | 1300 (38.0) | 1079 (42.0) | 2128 (39.6) | 251 (41.4) | 2379 (39.7) |
| Current | 382 (11.2) | 183 (7.1) | 515 (9.6) | 50 (8.3) | 565 (9.4) |
| **Physical activity, Hours per week** |  |  |  |  |  |
| Low | 1129 (33.0) | 634 (24.7) | 1564 (29.1) | 199 (32.8) | 1763 (29.5) |
| Moderate | 588 (17.2) | 379 (14.8) | 878 (16.3) | 89 (14.7) | 967 (16.2) |
| high | 1615 (47.3) | 1484 (57.8) | 2791 (51.9) | 308 (50.8) | 3099 (51.8) |
| **Diabetes** | 94 (2.8) | 63 (2.5) | 129 (2.4) | 28 (4.6) | 157 (2.6) |
| **BMI, kg/m<sup>2</sup>** | 25.9 (4.0) | 26.4 (3.8) | 26.1 (3.9) | 26.3 (4.1) | 26.1 (3.9) |
| **SBP, mmHg** | 121 (15.3) | 127 (17.5) | 123 (16.5) | 126 (16.6) | 123 (16.5) |
| **DBP, mmHg** | 76.4 (9.9) | 79.0 (11.2) | 77.6 (10.5) | 77.4 (10.4) | 77.5 (10.5) |
| **Glucose, mmol/L** | 5.2 (1.2) | 5.3 (1.2) | 5.2 (1.2) | 5.4 (1.6) | 5.2 (1.2) |
| **Total cholesterol, mmol/L** | 5.9 (1.0) | 5.9 (1.1) | 5.9 (1.0) | 6.1 (1.1) | 5.9 (1.1) |

Values are means (SDs) for continuous variables and numbers (percentages) for categorical variables

### Association between cardiac troponin I at baseline and incidence of dementia

5985 participants free of diagnosed cardiovascular disease and dementia at baseline (age range 45-69) were included in the analyses. Every doubling of cardiac troponin I was associated with 11% higher risk of dementia (HR=1.11, 95% CI: 1.04 to 1.19). The association did not differ between age group (≤60 vs >60 years) (P_interaction_=0.76) or sex (P_interaction_=0.32). Compared with those with cardiac troponin I below quantification limit (<2.5 ng/L), participants with cardiac troponin I >5.2 ng/L had 43% higher risk of dementia (HR=1.43, 95% CI: 1.13 to 1.80) (Table 2). We also plotted the association between cardiac troponin I and dementia incidence using natural cubic splines (Figure S1). The result was consistent with analysis with categorised cardiac troponin I group and showed continuously increased risk of dementia as cardiac troponin I increases. The increase in dementia risk tended to be smaller as troponin I level increased, however this was not a statistically significant effect (P for non-linearity=0.22).

**Table 2.** Association between high-sensitivity cardiac troponin I at baseline (1997-99) and incident dementia during 25 years of follow-up.

| **Cardiac troponin I, ng/L** | **No of cases/total No** | **HR (95% CI)** |
| --- | --- | --- |
| <2.5 | 158/2135 | 1.00 (reference) |
| 2.5-3.4 | 133/1282 | 1.22 (0.96,1.54) |
| 3.5-5.2 | 150/1281 | **1.34 (1.06,1.70)** |
| >5.2 | 165/1287 | **1.43 (1.13,1.80)** |

The model was adjusted for age, sex, ethnicity, education, occupational position, smoking status, alcohol consumption, physical activity, BMI, systolic blood pressure, diastolic blood pressure, glucose, and total cholesterol.

### Association between cardiac troponin I at baseline and cognitive trajectory

5895 participants free of diagnosed cardiovascular disease and dementia at baseline with one or more measurements of cognitive data were included in the analyses (90 participants without any cognitive data were excluded). Of them, 2508 (42.5%) had cognitive data at all six waves, 1102 (18.7%) at five waves, 652 (11.1%) at four waves, 555 (9.4%) at three waves, 500 (8.5%) at two waves, and 578 (9.8%) at only one wave. Cognitive trajectory differed by cardiac troponin I after adjusting for sex, ethnicity, education level, occupational position, birth cohort (continuous values: P_interaction_=0.07; grouped values: interaction P=0.0012). Participants with higher cardiac troponin I at baseline had a faster decline of cognitive function with age (Figure 1). Compared with participants with cardiac troponin I <2.5 ng/L, those with cardiac troponin I >5.2 ng/L had similar global cognitive z score at age 60, but had 0.10 (0.02 to 0.18) standard deviations lower global cognitive z score at age 80, and 0.19 (0.03 to 0.35) standard deviations lower score at age 90 (Table 3). Results were similar after further adjusting for behavioural and biomedical risk factors at baseline (Figure S2, Table S1).

**Figure 1.**
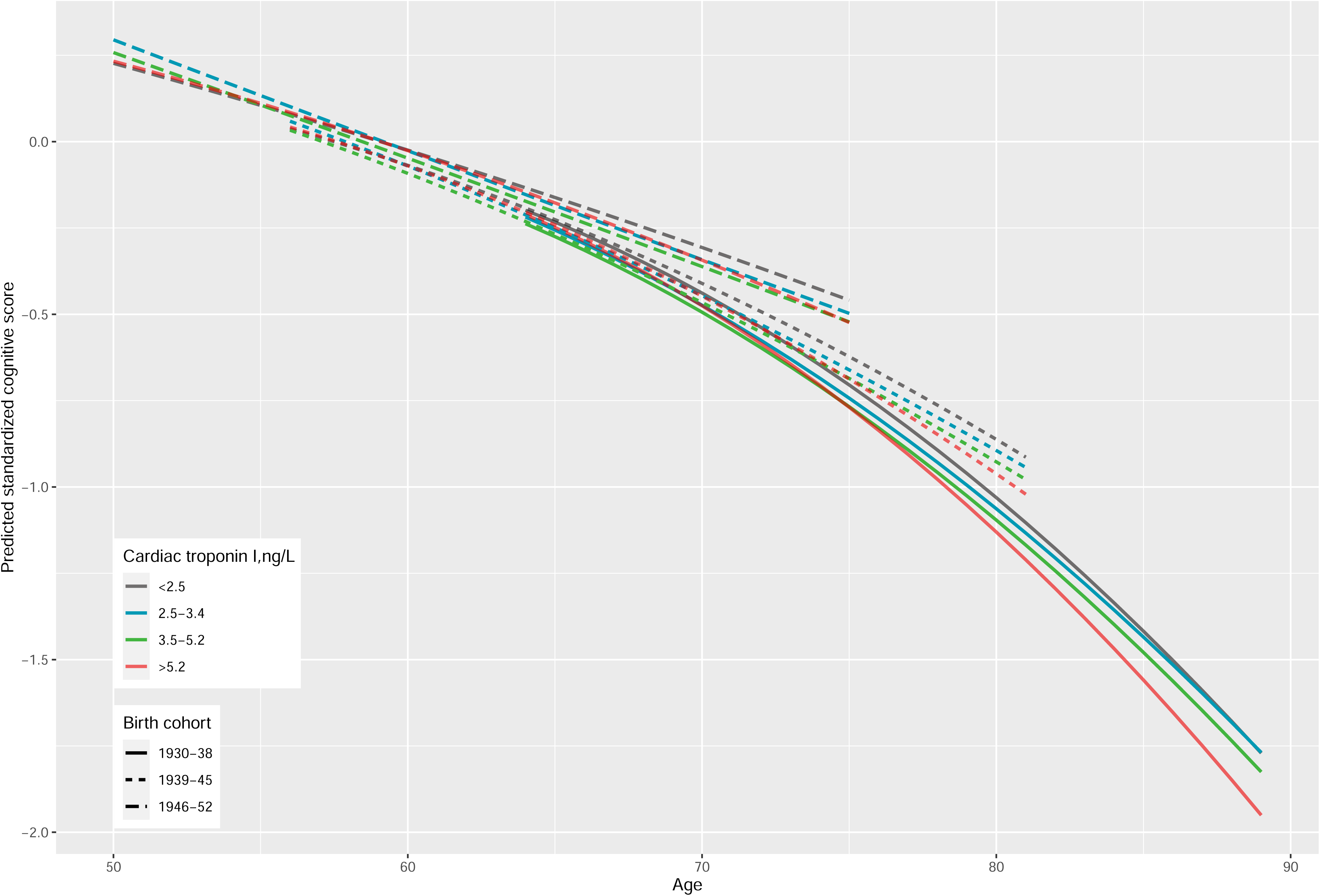
Cognitive trajectories from age 50 to 89 years by levels of high-sensitivity cardiac troponin I at baseline stratified by birth cohort. Predicted cognitive scores estimated from a mixed model (model terms: sex, ethnicity, education level, occupational position, birth cohort, age, age^2^, and interactions of all covariates with age and age^2^). P for difference in trajectory between different cardiac troponin levels: 0.0012.

**Table 3.**
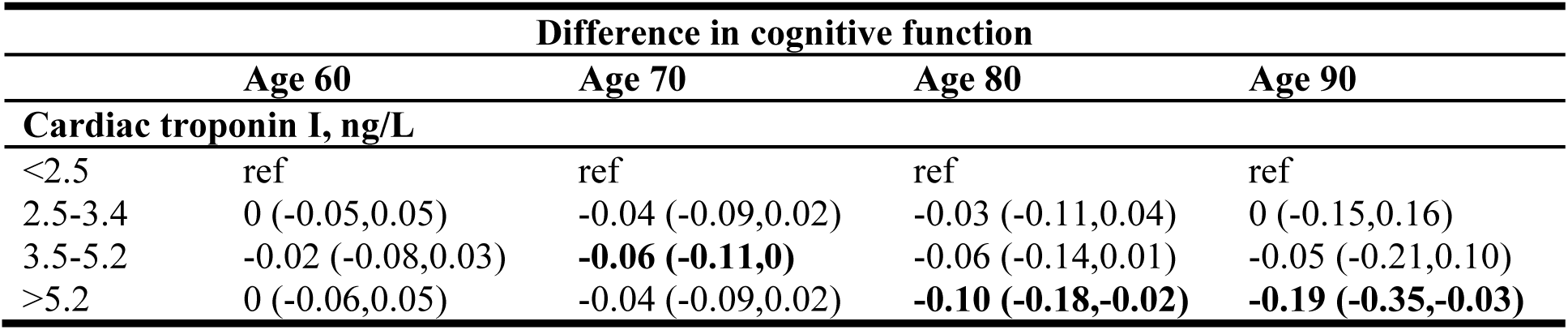
Association of high-sensitivity cardiac troponin I with cognitive performance at age 60, 70, 80, and 90 years.

### Backward trajectory of cardiac troponin I before dementia

Among the 695 incident dementia cases identified in the study sample, all were successfully matched to 4 controls and this led to a nested case-control sample of 3475 individuals. Both case and control groups had an increasing predicted cardiac troponin I level over time, which were not significantly different, though they tended to converge (interaction P=0.12) (Figure 2). Compared with controls, cases had consistently higher mean cardiac troponin I levels from 25 years to 7 years preceding dementia diagnosis (Table S2). Further matching by cardiovascular disease status showed similar but attenuated results (Figure S3, Table S3).

**Figure 2.**
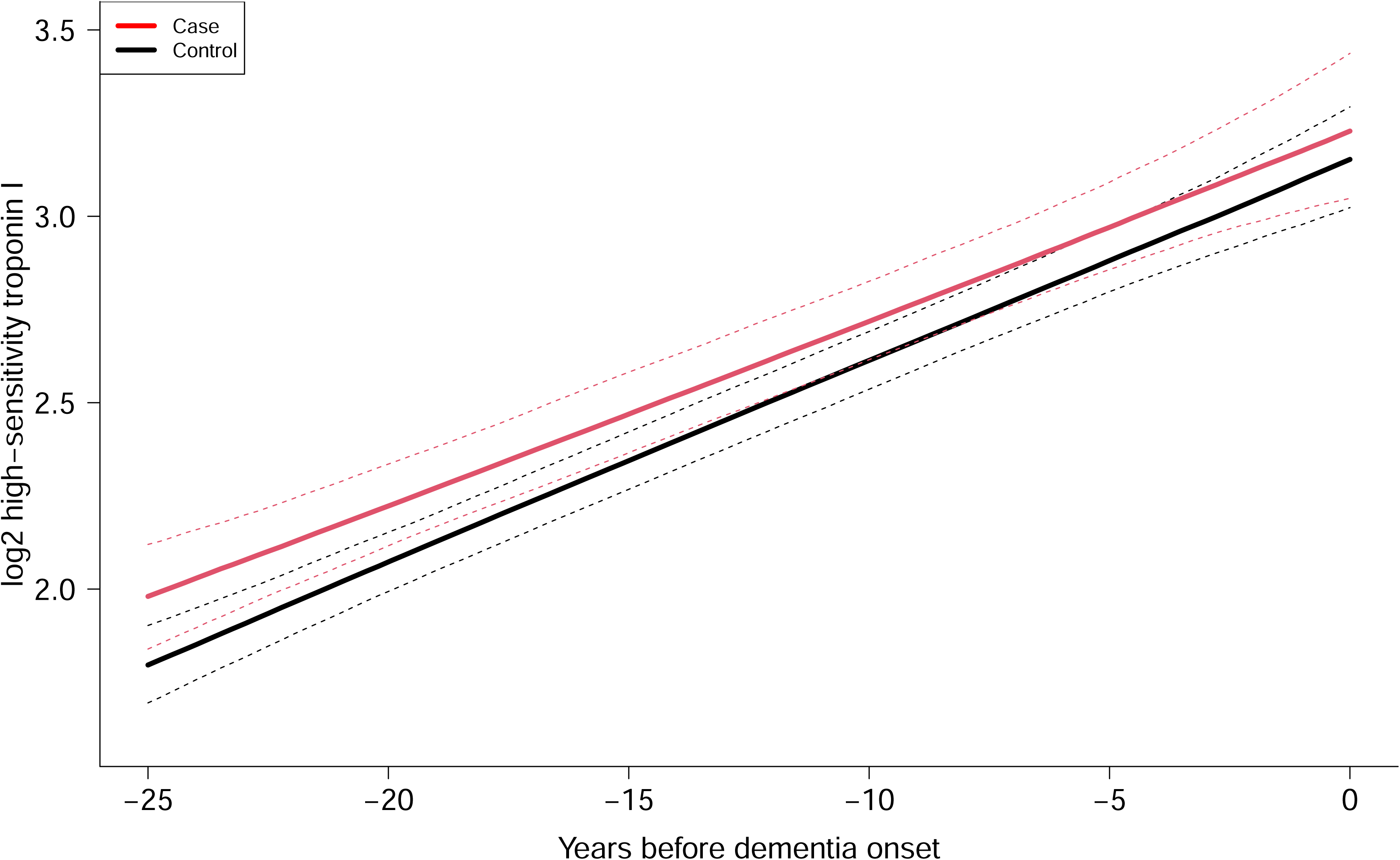
Averaged predicted trajectories of high-sensitivity cardiac troponin I (ng/L, log2 transformed) for 695 incident cases of dementia and 2780 matched controls over 25 years preceding diagnosis of dementia estimated from a latent process mixed model. Nested case-control design matching with age, sex and education level. Dashed lines represent 95% CIs. P for difference in trajectory between cases and controls: 0.12.

### Association between cardiac troponin I at baseline and brain volume

Of the 771 participants in the substudy, we excluded those without a cardiac troponin I measurement (n=129), or who had dementia or cardiovascular disease at baseline (n=1), producing an analytical sample of 641. One year older age was associated with 0.32% (95%CI: 0.28 to 0.35%) lower total brain volume, 0.23% (95%CI: 0.20 to 0.26%) lower grey matter volume, and 0.09% (95%CI: 0.06 to 0.11%) lower white matter volume. Higher cardiac troponin I level at baseline was associated with lower grey matter volume, but not with white matter volume. People with cardiac troponin I concentrations >5.2 ng/L had 0.63% (95%CI: 0.23 to 1.04%) lower grey matter volume and 18% (95%CI: 0 to 40%) increased risk of hippocampal atrophy compared with those with cardiac troponin I <2.5 ng/L, corresponding to an age effect of 2.7 and 3 years, respectively (Table 4). Cardiac troponin I concentration was not associated with white matter hyperintensities.

**Table 4.**
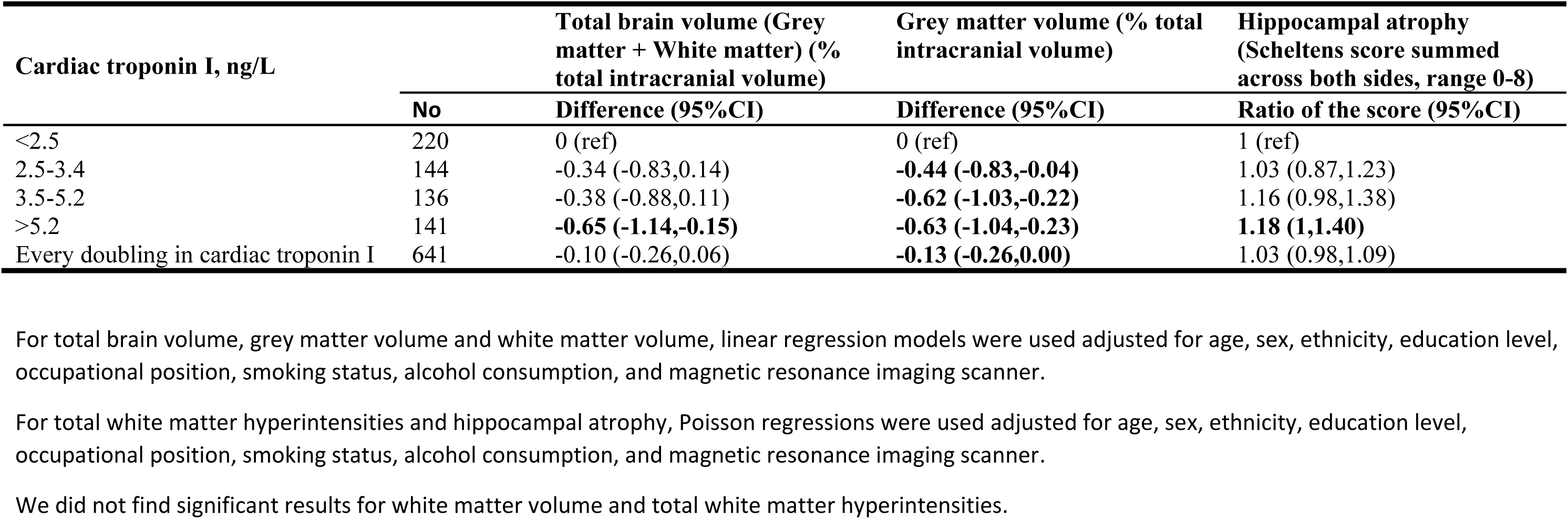
Association between high-sensitivity cardiac troponin I at phase 5 (1997-99) and structural brain volume 15 years later: Whitehall II imaging substudy (n=641)

## Discussion

Our longitudinal study suggests that subclinical myocardial injury in midlife, indicated by elevated cardiac troponin I levels, is associated with higher dementia risk. We have three key findings. First, people with higher cardiac troponin I concentrations at baseline had faster decline of cognition and were more likely to be diagnosed with dementia over a median 25 years of follow-up. Second, backward trajectory analysis showed that cardiac troponin I level was consistently higher in those who developed dementia compared to those who did not between 7 and 25 years before diagnosis. The length of time suggests that higher cardiac troponin I is unlikely to be due to prodromal changes, and it may be on the causal pathway. Third, in the MRI substudy, people with increased cardiac troponin I at the midlife baseline were more likely to have relatively low grey matter volume and hippocampal atrophy 15 years later. These three consistent findings involving repeated assessments with a high-sensitivity cardiac troponin I assay starting in midlife strengthen the evidence that myocardial injury may directly or indirectly contribute to the aetiology of dementia.

### Comparison with previous studies

Two published studies have examined the prospective association of cardiac troponin with incident dementia.^20^ The Atherosclerosis Risk in Communities (ARIC) study found that elevated levels of high-sensitivity cardiac troponin T measured at mean age of 62.5 years were associated with higher dementia incidence over 13 years of follow-up.^10^ The national FINRISK 1997 study reported that high-sensitivity cardiac troponin I measured at a mean age of 47.9 years was associated with incident dementia over 18 years of follow-up.^21^ Our study had longer follow-up and larger effect size compared with FINRISK using high-sensitivity troponin I (Abbott Architect), but a smaller effect size compared with the ARIC study using high-sensitivity troponin T (Roche Elecsys). These previous studies were based on a single measurement of cardiac troponin.

Our novel contribution to the evidence is based on repeated measurements of cardiac troponin I over a 15-year period which allowed the backward trajectory before dementia diagnosis to be modelled. We observed an increased cardiac troponin level occurred as early as 25 years before dementia diagnosis. Further, our characterisation of cognitive decline, based on six assessments of cognitive function over 25 years substantiates the interpretation that subclinical myocardial injury is associated with long-run processes in the midlife preclinical period.

A few prospective studies found higher cardiac troponin T concentration at baseline was associated with faster annual decline in global cognition. However, these studies either had cardiac troponin measured at older age (>65 years)^11,12,22^ or had few repeated measurements of cognitive function (<3 times),^23^ or had a short follow-up time (<5 years).^11,12^ Consequently, no previous study investigated the effect of the cardiac troponin at midlife on the subsequent long-term cognitive trajectory until late life. Our study measured cardiac troponin I at a mean age of 56 years and had up to 6 repeated measurements of cognitive function across 25 years and found that cognitive performance was not significantly different in the 60s and 70s, but started to diverge around age 80. This is consistent with the finding from the ARIC study that cardiac troponin T measured at a mean age of 63 years was not associated with 15-year change in cognitive function using only two repeated cognitive function measured 15 years apart.^23^

Few population-based studies have examined the association between cardiac troponin and structural brain changes, and cardiac troponin T assays were mostly used that have greater imprecision than high-sensitivity cardiac troponin I assays at the low concentrations observed in population studies.^24^ A cross-sectional study using data from the Maastricht study found that a higher cardiac troponin T concentration was associated with smaller grey matter volume and greater white matter hyperintensity among participants aged 60 and above.^25^ The ARIC study reported that individuals with higher cardiac troponin T was associated with MRI-defined silent brain infarcts and white matter lesions cross-sectionally and more white matter lesions progression on the follow-up MRI 11 years later.^26^ Our study used a high-sensitivity cardiac troponin I assay with consistent findings in regards to the smaller grey matter in those with higher cardiac troponin levels. A potential explanation is that grey matter is more susceptible to cerebral hypoperfusion caused by cardiac dysfunction as it has higher metabolic demand compared with white matter. Our study, however, did not find cardiac troponin to be associated with white matter hyperintensity. This is consistent with the previous evidence from the Whitehall II study that the life Simple 7 cardiovascular health score was not associated with white matter hyperintensities.^5^ A recent study using UK Biobank, however, found that higher Life Essential 8 cardiovascular health score was associated with lower white matter hyperintensity volume.^27^ This reflects that cardiac troponin may capture different aspects of cardiac dysfunction compared to cardiovascular health scores. Different methods used to measure white matter hyperintensity may also explain these inconsistent observations.

### Role of subclinical myocardial injury in dementia aetiology

Dementia and cardiovascular disease share risk factors such as hypertension and hyperlipidemia.^3,28–30^ These risk factors may contribute to the heart-brain connection by causing damage to vessels in both organs. Importantly, our analysis shows that higher cardiac troponin was associated with higher dementia risk independent of hypertension and hyperlipidaemia. Higher cardiac troponin is associated with increased risk of stroke,^31^ and stroke is known to double the risk of dementia.^32^ Interestingly, the effect was only slightly attenuated when adjusting for clinically diagnosed cardiovascular disease in our study, indicating other mechanisms beyond occlusion of major arteries, such as cerebral small vessel disease and cerebral hypoperfusion and hypoxia, may be involved.^3,33^ A previous review concluded that cardiac troponins are more consistently linked to vascular brain lesions than to neurodegenerative changes such as brain atrophy.^2^ However, we found that cardiac troponin was prospectively associated with brain atrophy. It is possible that vascular and neurodegenerative brain damage may overlap and develop in parallel^34^ and cardiac troponin may provide insights into both pathologic processes.

### Strengths and limitations

Our study, with a median follow-up of 25 years, is less susceptible to reverse causation. The development of dementia involves a long prodromal period, and neuropathological abnormality and change of biomarker levels can begin 15 to 20 years before clinical diagnosis of dementia. No other population-based studies have measured cardiac troponin level at midlife with a follow-up period of 20 years, which is critical to examine the association between subclinical myocardial injury in midlife and incident dementia.

Another novel strength is the incorporation of repeated measurements of high-sensitivity cardiac troponin I and modelling the backward trajectory of cardiac troponin before dementia diagnosis. Previous studies only focused cardiac troponin level at one time point, and its association with incident dementia depends on both when cardiac troponin was measured and the length of the follow-up period. This conventional study design cannot determine the length of time that cardiac troponin levels have been raised before dementia diagnosis.

Our results may underestimate the association as cardiac troponin is associated with mortality, although we would have picked up some of these people if dementia was recorded on the death certificate. Troponin I levels can be elevated due to other causes that cardiac muscle pathology such as intense exercise, but this would not be expected to increase the risk of dementia. The population in Whitehall II are predominantly white, and generalisation of our results to other ethnicity groups should be cautious. We used one cardiac troponin I assay, and validation of our results using other cardiac troponin assays is needed if measurement were to be used in clinical practice to inform dementia risk.

## Conclusion

Subclinical myocardial injury at midlife is associated with higher dementia incidence in later life. Measurement of cardiac troponin I using a high-sensitivity assay in midlife may be useful in the early identification of a population at risk of cognitive decline and dementia.

## Data Availability

Whitehall II data cannot be shared publicly because of constraints dictated by the study's ethics approval and IRB restrictions. The Whitehall II data are available for sharing within the scientific community. Researchers can apply for data access at https://www.ucl.ac.uk/psychiatry/research/mental-health-older-people/whitehall-ii/data-sharing.

## Acknowledgements

We thank all of the participating civil service departments and their welfare, personnel, and establishment officers; the British Occupational Health and Safety Agency; the British Council of Civil Service Unions; all participating civil servants in the Whitehall II study; and all members of the Whitehall II Study team at UCL and Oxford.

## Contributors

YC and EJB conceptualised and designed the study. YC and MS did the statistical analysis. YC wrote the first draft of the manuscript. All authors interpreted the data and critically revised the manuscript. All authors approved the final version of the manuscript for submission. YC and EJB are the guarantors. The corresponding author attests that all listed authors meet authorship criteria and that no others meeting the criteria have been omitted.

## Ethical approval

Written informed consent from participants and research ethics approvals were renewed at each contact. Ethical approval was from the University College London Hospital Committee on the Ethics of Human Research (reference number 85/0938).

## Transparency declaration

The manuscript is an honest, accurate, and transparent account of the study being reported. No important aspects of the study have been omitted. Any discrepancies from the study as originally planned (and, if relevant, registered) have been explained.

